# Cardiovascular and autonomic responses to transcutaneous spinal cord stimulation combined with activity-based therapy after chronic spinal cord injury: An exploratory study from the MACHINE trial

**DOI:** 10.64898/2026.08.11.26359978

**Authors:** Shane J.T. Balthazaar, Claire L. Shackleton, Alison M.M. Williams, Soshi Samejima, Raza N. Malik, Daniel D. Hodgkiss, Tom E. Nightingale, Rahul Sachdeva, Stacy L. Elliott, Michael J. Berger, Tania Lam, Andrei V. Krassioukov

## Abstract

**Objective:** To describe cardiovascular and autonomic responses to body weight-supported treadmill training (BWSTT) combined with active or sham transcutaneous spinal cord stimulation (TSCS) in individuals with chronic, motor-complete spinal cord injury (SCI).

**Design and setting:** Exploratory case series from randomized, sham-controlled clinical trial in a tertiary Rehabilitation Centre in Vancouver, Canada.

**Participants:** Eight adults with chronic (≥1 year post-injury) traumatic, motor-complete (American Spinal Injury Association Impairment Scale A-B) SCI at or above T6

**Interventions:** Participants were randomized to 12 weeks of BWSTT plus lumbosacral TSCS or BWSTT plus sham stimulation, delivered 3 sessions/week. TSCS was delivered at T11-L1 using 30 Hz stimulation with a 10 kHz carrier frequency. Five participants completed the intervention, and four completed full cardiovascular testing (TSCS n=2; sham n=2).

**Outcome measures:** Ambulatory blood pressure (BP) monitoring, participant-reported symptoms of AD and OH (via ADFSCI questionnaire), BP variability, orthostatic hemodynamics, echocardiography, electrocardiography (ECG)- and heart rate variability (HRV)-derived indices, and baroreflex function.

**Results:** Among complete cases, several cardiovascular indices changed over time, including reduced daytime hypotensive burden in TSCS participants, preserved nocturnal dipping, and small changes in stroke volume and ECG-derived variability indices; however, responses were heterogeneous and overlapped with Sham. Both TSCS and Sham participants showed reduced autonomic symptom scores, while low-frequency blood pressure variability responses during orthostatic stress were heterogeneous and did not indicate a pattern that was specific to a cohort.

**Conclusion:** Although preliminary, this exploratory complete-case analysis suggests that cardiovascular responses to BWSTT with active or sham TSCS are measurable but highly individualized after chronic motor-complete SCI. Given the small sample and overlapping Sham responses, findings are exploratory and larger trials are needed to determine whether TSCS augments cardiovascular autonomic adaptations to locomotor training.

## Introduction

Cardiovascular autonomic dysfunction is a common and clinically important consequence of spinal cord injury (SCI), particularly following cervical and upper thoracic injuries. Disruption of descending sympathetic pathways can contribute to persistent arterial hypotension,(1) orthostatic hypotension (OH),(2) autonomic dysreflexia (AD),(3) impaired circadian blood pressure (BP) regulation, and exaggerated BP instability during daily life.(4) These disturbances may be symptomatic, interfere with rehabilitation participation, and contribute to adverse cardiovascular consequences after SCI.(1,5) Despite this, cardiovascular autonomic outcomes have received relatively little attention in rehabilitation trials.

Activity-based therapies (ABT), including body-weight supported treadmill training (BWSTT), are most often used to engage spinal sensorimotor networks below the neurological level of injury and to improve motor or locomotor function.(6,7) However, the cyclic lower-limb movement combined with an upright postural challenge may influence the areas of the autonomic nervous system involved in cardiovascular regulation. Improvements in bladder, bowel and sexual function following locomotor-based rehabilitation suggest that spinal autonomic networks may remain modifiable after chronic injury.(8) Whether similar training can alter cardiovascular autonomic function, particularly in people with chronic motor-complete SCI, is less clear.

Transcutaneous spinal cord stimulation (TSCS) offers a non-invasive strategy to modulate spinal cord excitability. Mechanistic studies have shown that TSCS recruits posterior-root afferents and activates neural input structures that overlap with those engaged by epidural stimulation,(9) and cardiovascular effects may be mediated through somato-autonomic reflex pathways involving sympathetic preganglionic neurons and interneuron-based relays.(10) Early studies suggest that non-invasive spinal stimulation can acutely influence cardiovascular autonomic control after SCI, including orthostatic BP responses and AD.(11,12) However, responses are heterogeneous, and recent work indicates that TSCS may not uniformly improve autonomic regulation.(13) Whether repeated TSCS, delivered during locomotor training, induces sustained changes in cardiovascular control remains unknown.

Pairing TSCS with BWSTT may therefore provide a strategy to combine neuromodulation of spinal cord excitability with task-specific afferent input and repeated orthostatic cardiovascular challenge. Existing studies of TSCS combined with locomotor training have primarily evaluated walking function, motor performance and spasticity,(14,15) leaving the cardiovascular consequences of this paired intervention poorly characterized. Our Motor and Autonomic Concomitant Health Improvements with Neuromodulation and Exercise (MACHINE; NCT04726059) trial was designed as a single-blind, randomized, sham-controlled study of BWSTT paired with active TSCS or sham stimulation in individuals with chronic motor-complete SCI.(16) Here, we report the cardiovascular outcomes from the MACHINE trial, including ABPM and participant-reported symptoms of AD and OH, alongside exploratory measures of BP variability, orthostatic hemodynamics, echocardiography, electrocardiography (ECG)- and heart rate variability (HRV)-derived indices, and baroreflex function. As only a small number of participants completed the full cardiovascular battery, we interpret these data as exploratory rather than definitive.

## Methods

This study was approved by the University of British Columbia Clinical Research Ethics Board and Health Canada (Investigational Testing Authorization #336767) for use of the Transcutaneous Electrical Spinal Cord Neuromodulator (TESCoN) class II medical device and conducted at the International Collaboration on Repair Discoveries (ICORD), in the Blusson Spinal Cord Centre, University of British Columbia, Vancouver, Canada. Recruitment occurred from August 8, 2022, to May 28, 2024. All participants provided written informed consent before any study procedures.

### Participants

Participants were aged 18-60 years with chronic traumatic SCI ≥T6, AIS A or B,(17) ≥1 year post-injury were eligible. Participants were required to tolerate upright positioning for at least 30 minutes, have stable SCI-related management and medication dosage for at least 4 weeks, and be able to comply with all study visits. Key exclusion criteria included ventilator dependence, unmanaged depression, active substance abuse, intrathecal baclofen, severe acute medical issues, cardiovascular, respiratory, renal, bladder, or hepatic disease unrelated to SCI, implanted devices or metal near stimulation sites, seizure history, pregnancy, open wounds at stimulation sites, and other conditions judged by investigators to interfere with safe participation. Participants avoided caffeine, cannabis, meals, and vigorous activity up to three hours before testing.

### Randomization, allocation concealment, and blinding

Participants were randomized in a 1:1 ratio using a computer-generated simple randomization sequence. Clinical coordinators generated the randomization sequence, prepared sequentially numbered sealed envelopes, and enrolled participants. Envelopes were opened only after Pre-Intervention assessments were completed and the participant was ready for allocation. Participants were blinded to group allocation. Trainers were not blinded because they were required to deliver active or sham stimulation during training sessions. The cardiovascular outcome assessors remained blinded to allocation until completion of data processing.

### Study protocol and interventions

Participants completed BWSTT 3 times per week for 12 weeks, with a target of 36 sessions. Training was individualized according to participant tolerance and progressed by increasing walking duration, increasing treadmill speed, reducing rest time, and reducing body-weight support while maintaining appropriate gait kinematics. Training sessions were structured around a target of up to 45 minutes of walking. BP, heart rate, and rating of perceived exertion were monitored during training, while reducing body-weight support where possible and maintaining appropriate gait kinematics. Training exposure, stimulation amplitude, body-weight support, walking speed, perceived exertion, and walking distance were recorded at each session.(18)

For participants allocated to the TSCS cohort, stimulation was delivered using the TESCoN non-invasive spinal stimulation device.(19) Cathodal electrodes were positioned over the T11 and L1 spinous processes, with rectangular anodal electrodes placed bilaterally over the iliac crests. An asymmetric, charge-balanced monophasic waveform was delivered at 30 Hz with a 10 kHz carrier frequency. The waveform comprised a short-duration (1 ms), high-amplitude cathodic leading phase followed by a longer-duration (10 ms), lower-amplitude anodic phase with equal total charge to the cathodic phase, ensuring net zero charge delivery. TSCS intensity was set based on comfort, with all participants tolerating the maximum 130 mA. Stimulation was discontinued in the event of adverse events (e.g., AD).

Participants allocated to Sham underwent identical training and electrode placement procedures. To maintain blinding, stimulation was briefly increased to sensory threshold and then turned off for the remainder of the session. This sham approach, adapted from prior balance and walking studies,(20) and designed to maintain blinding by simulating the sensation of stimulation without delivering therapeutic effects.

### Outcomes

Prespecified cardiovascular outcomes(16) included 24-hour ambulatory BP monitoring (ABPM) measures of daytime and nighttime BP, and frequency and severity of AD and OH using the Autonomic Dysfunction Following Spinal Cord Injury (ADFSCI) questionnaire. Additional cardiovascular assessments, including BPV, beat-to-beat orthostatic hemodynamics, echocardiographic cardiac structure and function, and electrocardiographic (ECG) interval variability, were collected to characterize individual responses pre- and post-intervention assessment visits and are presented as exploratory.

#### Blood pressure instability

Twenty-four-hour ABPM was used to assess BP regulation. Measurements were collected using the Meditech Card(X)plore device (Meditech, Budapest, Hungary), using a well-established protocol.(21) An appropriately sized cuff was placed on the non-dominant arm, with measurements every 15 minutes during participant-defined daytime and hourly during nighttime periods based on individual sleep schedules. Participants maintained activity logs noting catheterizations, meals, posture changes, physical activity, and BP symptoms. Data were analyzed offline using CardioVisions software (Meditech, Budapest, Hungary).

Outcomes included average daytime and nighttime BP, frequency of hypertensive “AD events”, and hypotensive episodes (systolic BP [SBP] <100 mmHg and diastolic BP [DBP]<70 mmHg, excluding nighttime). AD events were defined using an absolute SBP threshold >150 mmHg,(22) consistent with clinical practice. Nocturnal dipping was calculated using the averaged lowest nighttime values relative to median daytime BP. Time-domain BPV metrics (i.e., standard deviation (SD) of 24-hour BP readings, the coefficient of variation (CoV), average real variability (ARV), and variability independent of the mean (VIM) were derived to assess free-living BP regulation and ABT-related changes.(23)

#### Cardiac structure and function

Transthoracic echocardiography assessed cardiac structure and function using a 2.5 MHz phased-array transducer (Vivid 7, GE Medical, Norway) with participants in the left lateral decubitus position following five minutes of rest. Image acquisition followed guidelines set by the American Society of Echocardiography,(24) (parasternal and apical views), captured at end-tidal expiration, and analyzed offline using EchoPAC software (GE Healthcare, Horton, Norway). Left ventricular structure and global systolic and diastolic function were calculated as the average of three cardiac cycles.(25,26)

#### Continuous electrocardiogram

Resting lead II ECG was continuously recorded at 1000 Hz (Powerlab 16/35, AD Instruments, Colorado Springs, United States) via commercially available software (LabChart 8, AD Instruments, Colorado Springs, United States) with ECG Analysis Module (version 2.4) during a 5-minute resting period prior to other assessment testing.(27) Automated analysis identified P-wave, PR, QRS onset, T_peak_, and T_end_; QT and QTc (Bazett) were derived. All interval detections were visually inspected and manually corrected where necessary. Segments containing ectopy, motion artifact, or signal instability were excluded prior to spectral and variability analyses. These electrophysiological indices reflect variability in ventricular repolarization; however, clinical arrhythmic risk cannot be inferred from these measures in a small cohort. Variability for ECG intervals of interest (P-wave duration, PR interval, and T_peak_-T_end_) was calculated as the variance of the longest artifact-free segment and utilizing an autoregressive spectral analysis using a custom script developed in R (version 4.5.2, R Foundation for Statistical Computing, Vienna, Austria). QTVI was calculated as log10 normalized QT variance over RR variance values overall.(27)

#### Heart rate variability

HRV was assessed using LabChart with the HRV Analysis Module (version 2.0.3).(28) R-R intervals from 5-minute ECG recordings were visually inspected and corrected for artifacts or ectopic beats (600-1400 ms; complexity 0.7-1.5). Frequency-domain HRV analysis used the Lomb-Scargle periodogram, a spectral estimation method suited for unevenly sampled data, such as the R-R Interval time series. Power spectral density was calculated for LF (0.04-0.15 Hz) and HF (0.15-0.4 Hz) bands, with normalized LF and HF power representing autonomic modulation, across the analyzed recording segments.

#### Baroreflex Sensitivity and Effectiveness Analysis

Spontaneous Cardiovagal Baroreflex Sensitivity (cvBRS) and Baroreflex Effectiveness Index (BEI) were calculated from beat-by-beat SBP and R-R interval data using a custom script developed in R (version 4.5.2, R Foundation for Statistical Computing, Vienna, Austria). SBP and R-R intervals were analyzed to identify spontaneous baroreflex sequences of ≥3 beats with progressive increases (up) or decreases (down), requiring ≥1DmmHg SBP and ≥1 ms R-R change.(29) For each valid sequence, simple linear regression was used to relate SBP to the R-R Interval. Sequences with a correlation coefficient (*r*) ≥ 0.85 were used.(29) The average slope across pooled sequences defined the spontaneous cvBRS index (ms/mmHg), with separate up- and down-sequence values calculated. BEI quantified the proportion of SBP ramps followed by corresponding R-R responses, calculated separately for upward and downward ramps, reflecting baroreflex engagement independent of response magnitude.

#### Orthostatic intolerance: Head-up tilt test and hemodynamics

Continuous beat-to-beat hemodynamics (systolic/diastolic BP, mean arterial pressure, heart rate [HR]) were recorded using finger photoplethysmography with ECG (Finapres Nova, Finapres Medical Systems BV, Netherlands), with episodic brachial BP calibration every minute (Dinamap Pro, GE Healthcare, Chicago, United States) via LabChart. Following 10 minutes supine rest, participants underwent a 60° head-up tilt for 10 minutes and returned to supine for 5 minutes of recovery. Data were recorded and analyzed offline via LabChart to identify postural BP changes. In addition, stroke volume (SV), cardiac output (Q), maximum rate of pressure increase in systole (dP/dt_max_; indicating cardiac contractility), and total peripheral resistance (TPR) were derived from finger BP waveforms using the validated Modelflow® method.(30)

Digitized signals were analyzed at 1 ms resolution in MATLAB R2025b, with ectopic beats corrected by linear interpolation. The final 60 seconds of pre-tilt supine data were used for Pre-Intervention BP and HR analyses. BPV was assessed using continuous wavelet transform (analytic Morse wavelet) to generate scalograms representing time-frequency power distributions. Total wavelet power within selected frequency bands was calculated as an index of BP variability. Low-frequency (LF; 0.04-0.15 Hz) BPV, reflecting sympathetic vascular activity, was quantified as the difference between mean LF power during the 5-minute Pre-Intervention and the 5-minute period following tilt onset.(31)

#### Severity of cardiovascular dysfunction

The ADFSCI questionnaire was used to assess participant-reported symptoms of BP dysregulation. Sections 3 and 4 evaluated symptoms of AD and OH, respectively, using a 5-point Likert scale (0–4). AD scores were calculated as the mean of 18 symptom items, and OH scores as the mean of 32 items across functional contexts. Higher scores indicate greater symptom burden.(32)

### Statistical analysis

We analyzed participants who completed all relevant cardiovascular assessments. The MACHINE study was designed as a randomized sham-controlled trial; however, due to the sample size, cardiovascular outcomes were analyzed descriptively as an exploratory case series. Individual median change scores were calculated for each outcome variable (Δ = Post-Pre). Participants who completed training exposure but lacked post-intervention cardiovascular testing were excluded from physiological outcome analyses. Continuous physiological data (e.g., BP, HR, HRV) were summarized as supine means over 5 minutes and HUTT responses defined as maximal or minimal deviations from supine. Analyses were conducted in R (Version 4.5.2, R Foundation for Statistical Computing, Vienna, Austria). Graphical representations were created in R Studio and Adobe Illustrator (Version 30.0, Adobe Inc., 2025, San Jose, CA, USA).

To explore whether cardiovascular changes may have been influenced by participation in BWSTT rather than stimulation allocation, we conducted an exploratory pooled complete-case analysis in participants who completed the full cardiovascular testing battery. Directionally consistent changes across TSCS and Sham participants were interpreted as compatible with a shared training, repeated-testing, or study-period effect, whereas divergent responses were interpreted as not attributable to BWSTT alone. This analysis was descriptive only, and no inferential testing was performed because of the small sample size and absence of a no-training control group.

### Safety

Adverse events and cardiovascular safety events were monitored during training and testing. BP elevations were managed according to a predefined safety approach that included pausing or modifying training, reducing stimulation intensity, checking common AD triggers, or terminating the session when needed.

## Results

A total of 8 individuals were enrolled in the study. Five participants completed the full study protocol (n = 3, TSCS; n = 2, Sham; Figure 1) with only four participants completing the full cardiovascular testing battery (TSCS n = 2; Sham n = 2). One Sham participant initiated training but discontinued before post-intervention cardiovascular testing because of unrelated surgery. Detailed demographics and injury characteristics are presented in Table 1. Detailed measures at Pre-Intervention and Post-Intervention can be found in Supplementary Table 1.

**Figure 1.**
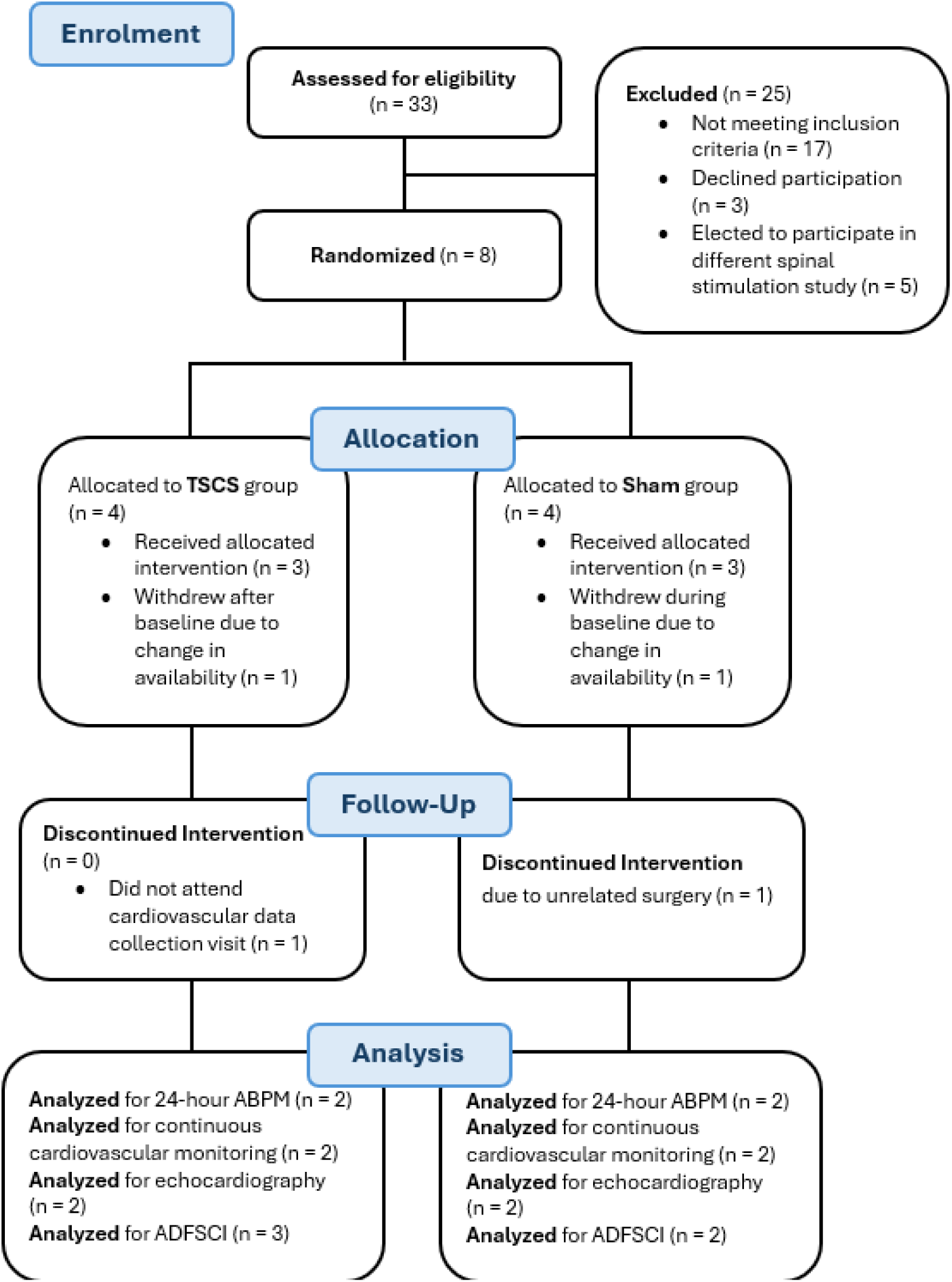
CONSORT Diagram.

**Table 1.** Participant demographics, injury characteristics, stimulation parameters, training dose.

| Participant | Cohort | Sex | Age Range (years) | NLI | AIS | TSI (years) | Mode, Pulse Width<br>Frequency<br>Average Maximum Amplitude, range | Sessions completed, n/N | BWS start → end, % body weight | Average Maximum BWSTT Walking Speed / session (km/h) | Average Maximum RPE / session | Total walking distance (m) / session, range |
| --- | --- | --- | --- | --- | --- | --- | --- | --- | --- | --- | --- | --- |
| 1 | TSCS | M | 40-45 | T5 | A | 20 | Monophasic, 1ms<br>30 Hz, 10 kHz (carrier)<br>85-130 mA | 36/36 | 41 → 41 | 2.7 | 9 | 1437-1649 |
| 2 | TSCS | M | 40-45 | T6 | A | 2 | Monophasic, 1ms<br>30 Hz, 10 kHz (carrier)<br>85-130 mA | 36/36 | 28 → 21 | 2.1 | 11 | 1060-1658 |
| 3 | Sham | M | 40-45 | C5 | A | 8 | - | 36/36 | 75 → 75 | 1.4 | 9 | 396-1113 |
| 4 | Sham | M | 35-39 | C7 | B | 30 | - | 34/36 | 65 → 59 | 1.9 | 11 | 778-1615 |
| 5 | TSCS | F | 35-39 | T1 | A | 33 | Monophasic, 1ms<br>30 Hz, 10 kHz (carrier)<br>54-69 mA | 28/36 | 71 → 71 | 2.0 | 13 | 989-1556 |

A, motor and sensory complete injury; AIS, American Spinal Injury Association Impairment Scale; B, motor complete and sensory incomplete injury; BWSTT, body-weight supported treadmill training; F, female; M, male; NLI, neurological level of injury; TSCS, transcutaneous spinal cord stimulation combined with locomotor therapy; TSI, time since injury in years

### 24-hour blood pressure, variability, and autonomic event frequency

**Figure 2.**
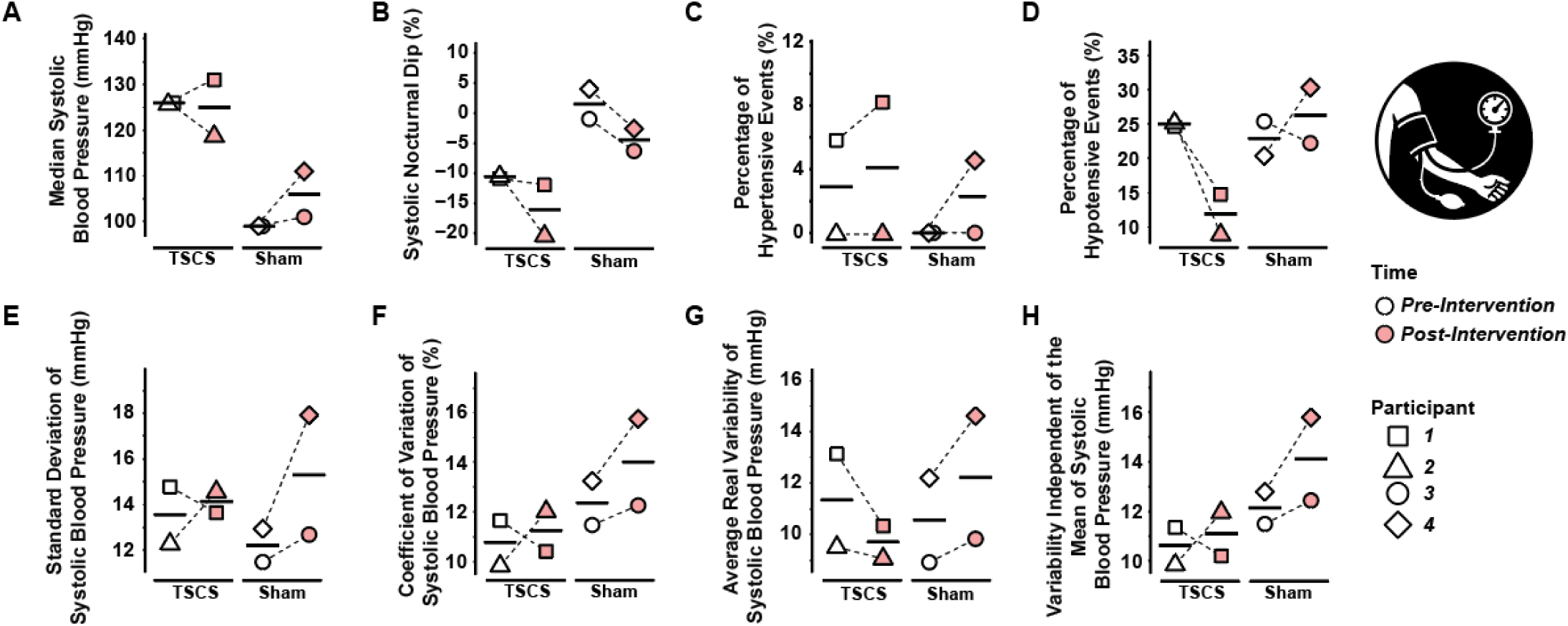
24-hour ambulatory blood pressure monitoring at Pre-Intervention and Post-Intervention. Individual data points (shapes) with horizontal bars indicating cohort medians. (**A**) Median systolic blood pressure (SBP) over 24 hours, (**B**) Systolic nocturnal dip to measure how much nighttime SBP decreases, (**C**) Percentage of hypertensive events (SBP > 150 mmHg), (**D**) Percentage of hypotensive events (SBP < 100 mmHg and diastolic BP < 70 mmHg), (**E**) Standard Deviation of SBP, (**F**) Coefficient of Variation of SBP, (**G**) Average Real Variability of SBP, (**H**) Variability Independent of the Mean of SBP.

#### Systolic Blood Pressure and Nocturnal Dipping

At Pre-Intervention, TSCS participants showed higher 24-hour median SBP than Sham (126 vs. 99 mmHg; Figure 2A). Sham demonstrated impaired nocturnal dipping (Δ +1.5%), whereas TSCS exhibited preserved dipping (Δ −10.6%). By Post-Intervention, both cohorts improved similarly (TSCS: Δ −5.45%; Sham: Δ −5.95%; Figure 2B), reflecting directional change over time, without evidence of a TSCS-specific effect.

#### Autonomic Dysreflexia and Daytime Hypotensive Events

AD episodes were present at Pre-Intervention in TSCS (2.9%) but absent in Sham. AD burden increased in both cohorts, more in Sham (Δ +2.25%) than TSCS (Δ +1.20%; Figure 2C). Daytime hypotension decreased with TSCS (Δ −13.05%) but showed inconsistent changes with Sham (Δ +3.35%; Figure 2D). Daytime hypotensive events decreased in both TSCS participants after training, with the median falling from 25.0% to 12.0%. In contrast, the Sham group showed divergent individual responses: one participant had fewer daytime hypotensive events at Post-Intervention, whereas the other had more. The Sham median therefore increased from 22.9% to 26.3%.

#### Blood Pressure Variability

The TSCS cohort showed reduced ARV-SBP (Δ −1.63 mmHg), with other variability indices showed mixed individual responses (Figure 2E-H); one participant had lower SD-SBP, CoV-SBP, and VIM-SBP at Post-Intervention, whereas the other had higher values. As a result, median SD-SBP, CoV-SBP, and VIM-SBP showed a modest change. In the Sham cohort, both participants showed higher SD-SBP, CoV-SBP, ARV-SBP, and VIM-SBP at Post-Intervention, though this is a descriptive participant-level observation due to sample size, but does not support an improvement in BPV after Sham training alone.

### Cardiac alterations following 12 weeks of TSCS and Sham interventions

**Figure 3.**
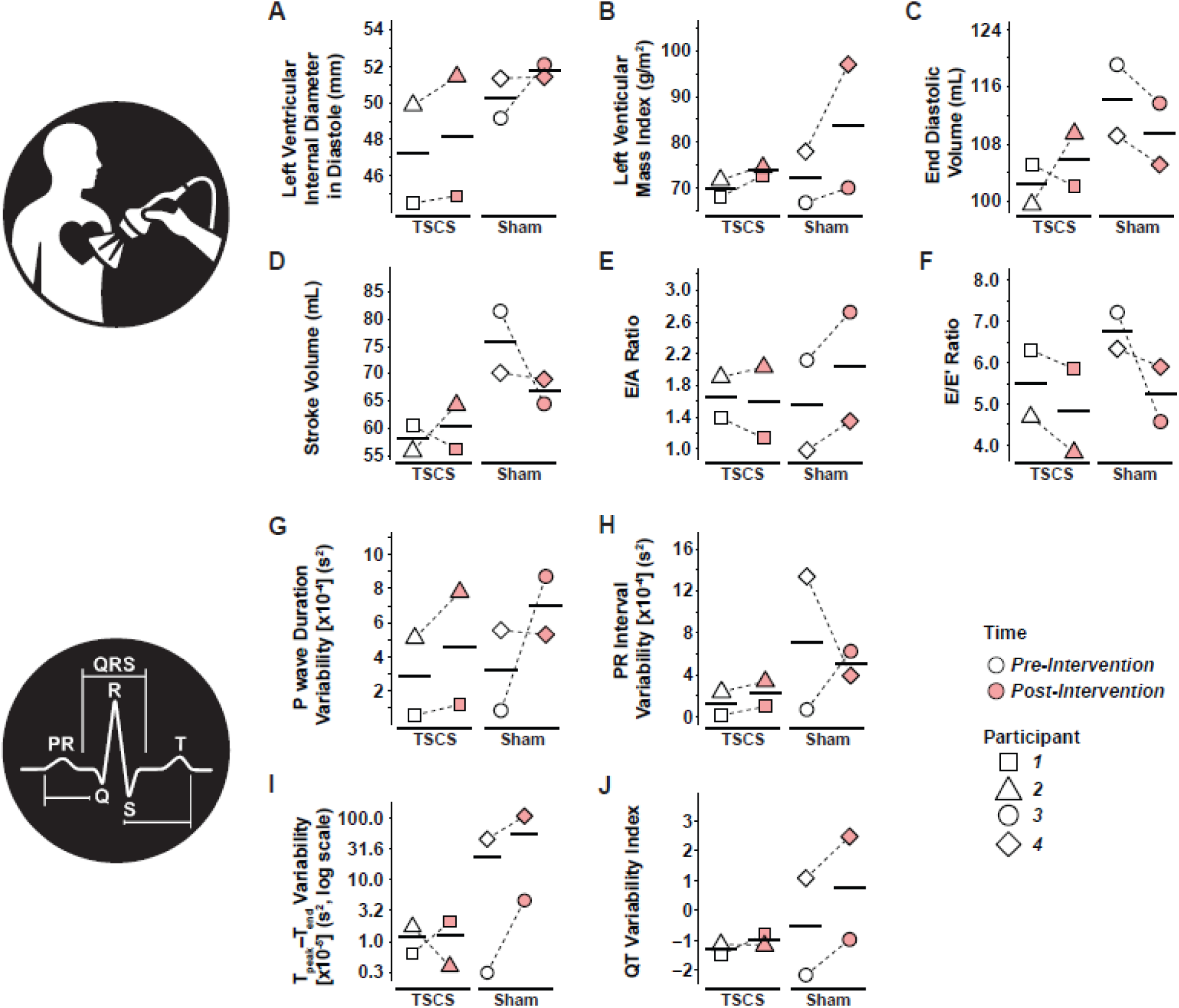
Echocardiographic indices of cardiac structure and function and ECG interval variability at Pre-Intervention and Post-Intervention. Individual data points (shapes) with horizontal bars indicating cohort medians. (**A**) Left ventricular (LV) internal diameter in diastole to measure chamber size, (**B**) LV mass index to help determine how thick the LV is with associated risks, (**C**) End diastolic volume to show how full the LV is before it pumps, (**D**) Stroke volume to measure how much blood the LV ejects with each heartbeat, (**E**) E/A Ratio to compare early passive filling in diastole to atrial contraction-driven filling, (**F**) E/E’ Ratio estimates LV filling pressure by comparing transmitral inflow velocity to early mitral annular tissue velocity (how well the LV relaxes). (**G**) P wave duration variability measuring the beat-to-beat changes in how long atrial depolarization takes, (**H**) PR interval variability measuring beat-to-beat changes in conduction time from the atria to the ventricles, (**I**) T_peak_-T_end_ variability measuring beat-to-beat changes in ventricular repolarization, (**J**) QT Variability Index quantifies how unstable the QT interval is compared to heart-rate variability for risk of arrhythmias.

For Pre-Intervention echocardiographic indices, left ventricular internal diameter in diastole (LVIDd) and left ventricular mass index (LVMi) were comparable between cohorts. Following the intervention in both groups, LVIDd increased slightly (TSCS +0.98□mm; Sham +1.51□mm; Figure 3A) and LVMi increased modestly (TSCS +3.74□g/m^2^; Sham +11.17□g/m^2^; Figure 3B). Stroke volume decreased in Sham (Δ −9.09□mL) but increased in TSCS (Δ +3.08□mL; Figure 3D). Diastolic indices showed small shifts: TSCS E/A decreased from 1.65 to 1.59 (Δ −0.06) and E/E’ from 5.51 to 4.85 (Δ −0.66); Sham E/A increased from 1.55 to 2.04 (Δ +0.49) with E/E’ decreasing from 6.78 to 5.23 (Δ −1.55; Figure 3E-F). From an ECG interval variability perspective, Pre-Intervention conduction intervals were similar. At Post-Intervention, R-R interval increased in TSCS (Δ +261.9□ms) and remained unchanged in Sham (Δ +0.5□ms). P-wave duration increased in TSCS (Δ +5.0□ms) and decreased in Sham (Δ −7.9□ms), with minimal PR changes (Figure 3G-H). QRS duration declined in TSCS (Δ −9.9□ms) and Sham (Δ −3.7□ms). Raw QT changed divergently (TSCS Δ +26.6□ms; Sham Δ −63.0□ms), but QTc decreased in both (TSCS Δ −17.4□ms; Sham Δ −54.3□ms). T_peak_-T_end_ remained stable in TSCS (Δ −0.1□ms) and declined slightly in Sham (Δ −5.6□ms; Figure 3C). QTVI remained negative in TSCS (−0.80 to −1.14), whereas Sham showed heterogeneity including an outlier (2.46; Figure 3J).

### Autonomic and Hemodynamic Responses to Orthostatic Stress

**Figure 4.**
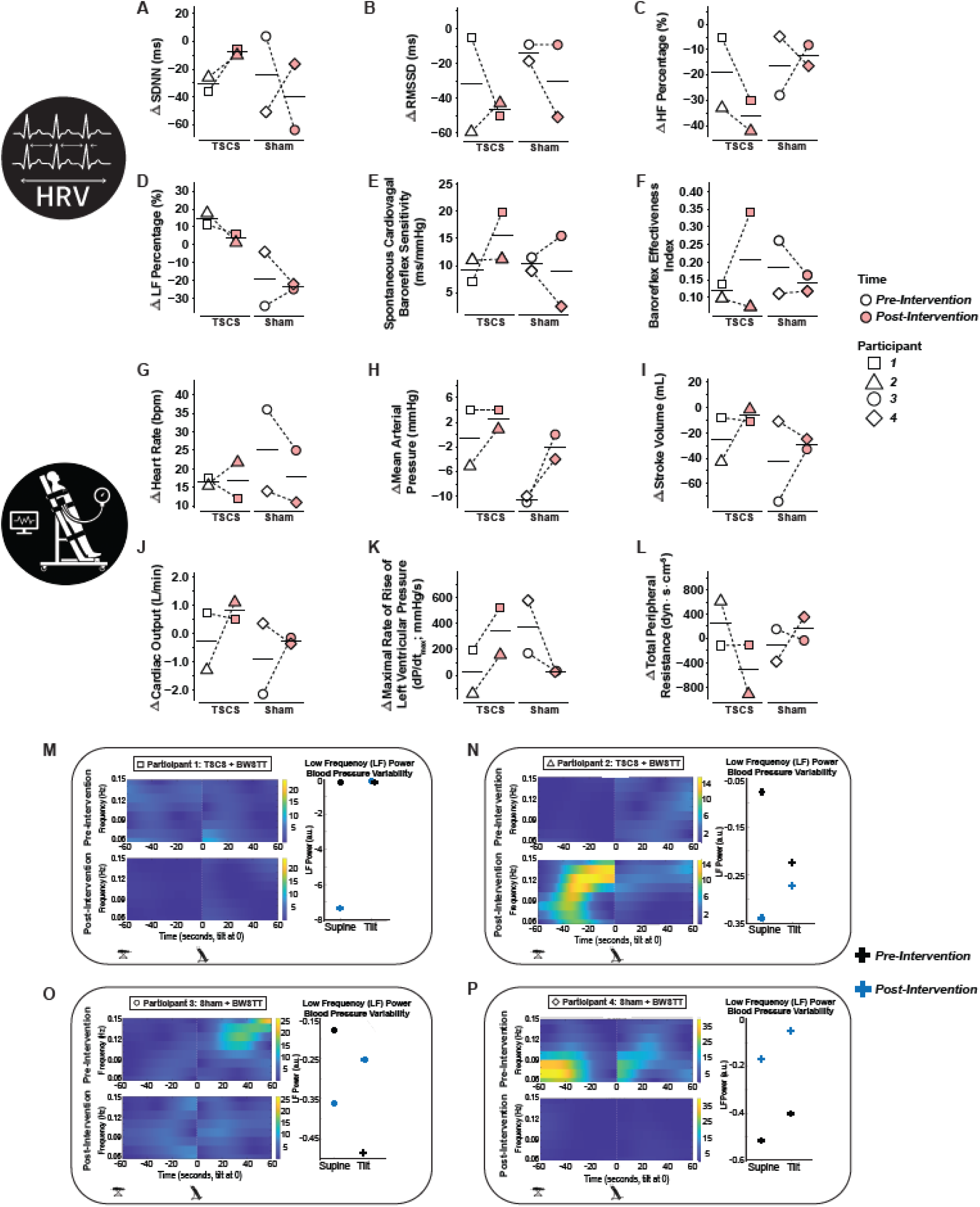
Heart rate variability, baroreflex, hemodynamic, and blood pressure variability responses to head-up tilt at Pre-Intervention and Post-Intervention. Individual data points (shapes) with horizontal bars indicating cohort medians from supine to tilted position following 10 minutes of rest. (**A**) Change (Δ) in standard deviation of normal-to-normal intervals (SDNN); (**B**) Δ root mean square of successive differences (RMSSD); (**C**) Δ high-frequency (HF) power (%); (**D**) Δ low-frequency (LF) power (%); (**E**) spontaneous cardiovagal baroreflex sensitivity; and (**F**) baroreflex effectiveness index during HUTT. (**G**) Δ heart rate; (**H**) Δ mean arterial pressure; (**I**) Δ stroke volume; (**J**) Δ cardiac output; (**K**) Δ maximal rate of rise of left ventricular pressure (dP/dt_max_); and (**L**) Δ total peripheral resistance from ahead-up tilt test (HUTT). (**M-P**) Representative continuous wavelet time-frequency scalograms of systolic blood pressure variability and corresponding low-frequency (LF) blood pressure variability during HUTT. Panels show Pre-Intervention (top) and Post-Intervention (bottom) recordings for each participant.

#### Autonomic Cardiac Regulation (HRV and Baroreflex)

Pre-Intervention HRV indices were elevated, with TSCS showing higher supine RMSSD and HF power. At Post-Intervention, supine HRV improved in both cohorts, with TSCS demonstrating increases in SDNN (Figure 4A), RMSSD (Figure 4B), HF (Figure 4C), and LF power (Figure 4D), whereas Sham improved mainly in time-domain measures with slight HF reduction (Supplementary Table 1). During tilt, TSCS exhibited higher HF and LF power at Pre-Intervention; post-intervention SDNN increased (Δ +36.37 ms) while HF (Δ −14.53%) and LF power (Δ −6.77%) declined; Sham changes were smaller. Pre-Intervention cvBRS (Figure 4E) and BEI (Figure 4F) were low and similar between cohorts. Following TSCS, one participant showed increased sequence frequency (6.98 to 19.76) and BEI (0.14 to 0.34), primarily via down-sequences; Sham responses were inconsistent.

#### Orthostatic Hemodynamics

At Pre-Intervention, TSCS participants demonstrated slightly higher HR (Figure 4G) and higher HUTT SBP, DBP, and MAP compared with Sham (SBP: 123 vs. 103DmmHg; DBP: 75 vs. 58DmmHg; MAP: 87 vs. 73DmmHg; Figure 4H). SV (Figure 4I) and Q (Figure 4J) were comparable, while dP/dt_max_ (Figure 4K) and TPR (Figure 4L) were higher in TSCS (Supplementary Table 1). At Post-Intervention, TSCS showed larger reductions in SBP (Δ −6DmmHg) and MAP (Δ −2DmmHg) with minimal DBP change, whereas Sham demonstrated smaller SBP reductions (Δ −4DmmHg) but increases in DBP (Δ +8DmmHg) and MAP (Δ +5DmmHg). HR decreased in both cohorts (TSCS Δ −15Dbpm; Sham Δ −11Dbpm). SV increased modestly (TSCS Δ +7 mL; Sham Δ +1 mL), Q decreased (TSCS Δ −0.76DL/min; Sham Δ −0.28DL/min), and dP/dt_max_ declined more in TSCS (Δ −347 vs. −214DmmHg/s). TPR increased in both cohorts, greater in Sham (Δ +565 vs. +29 dyn·s·cm^-5^).

#### Frequency-Domain Blood Pressure Variability

LF BPV responses to HUTT were heterogeneous across participants. Supine-to-tilt ΔLF responses at Pre-Intervention were 0.0 and −0.14 a.u. in the TSCS cohort, and −0.32 and +0.11 a.u. in the Sham cohort. At Post-Intervention, ΔLF responses were +7.3 and +0.07 a.u. in TSCS, and +0.11 and +0.13 a.u. in Sham. Visual inspection of the wavelet scalograms suggested less temporally clustered LF oscillatory activity during orthostatic stress in Participants 3 and 4; however, responses were heterogeneous across participants and point-estimate LF BPV did not indicate a consistent intervention- or cohort-specific effect (Figure 4M-P).

### Symptom burden of autonomic dysfunction in TSCS and Sham cohorts from ADFSCI

**Table 2.** AD and OH Scores Change from Pre-Intervention to Post-Intervention measures with standard autonomic dysfunctions questionnaire (ADFSCI)

| Participant | Cohort | AD<br>Symptom<br>Score<br>Pre-Intervention | AD<br>Symptom<br>Score<br>Post-Intervention | ΔAD | OH<br>Symptom<br>Score<br>Pre-Intervention | OH<br>Symptom<br>Score<br>Post-Intervention | ΔOH |
| --- | --- | --- | --- | --- | --- | --- | --- |
| 1 | TSCS | 1.17 | 1.00 | -0.17 | 0.05 | 0 | -0.05 |
| 2 | TSCS | 0 | 0 | 0 | 0 | 0 | 0 |
| 3 | Sham | 1.00 | 0.50 | -0.50 | 1.11 | 0.95 | -0.16 |
| 4 | Sham | 0.33 | 0.17 | -0.16 | 0.32 | 0.10 | -0.22 |
| 5 | TSCS | 2.00 | 1.50 | -0.50 | 1.05 | 0.70 | -0.35 |

Individual participant pre- and post-intervention scores for autonomic dysreflexia (AD) and orthostatic hypotension (OH), including change scores ( $\Delta$ = Post- Pre). Higher scores reflect greater symptom burden. ADFSCI, autonomic dysfunctions following spinal cord injury.

Pre-Intervention AD symptom severity was higher in the TSCS participants (1.06) than in Sham (0.67). Both cohorts showed reductions following the intervention (TSCS Δ −0.22; Sham Δ −0.33). Pre-Intervention OH scores were elevated in the TSCS participants (0.72 vs. 0.37 in Sham). Both cohorts improved (TSCS Δ −0.13; Sham Δ −0.19).

#### Exploratory pooled BWSTT complete-case analysis

We pooled the four participants who completed the full cardiovascular testing battery, irrespective of allocation. Nocturnal dipping improved consistently across all four participants, with a pooled median change of −5.95%, suggesting that this outcome may reflect a shared intervention-period effect rather than a TSCS-specific response. E/E’ also decreased in all four participants, and QTc decreased in three of four participants, indicating directionally consistent changes in selected cardiac indices. However, daytime hypotensive burden, ARV-SBP, resting stroke volume, and baroreflex indices showed heterogeneous responses between TSCS and Sham participants. These findings do not support attributing all cardiovascular changes to BWSTT alone and suggest that these specific outcomes may be compatible with a BWSTT-associated (or non-study related) effect, while other outcomes remain potentially stimulation-modulated (or individually variable).

## Discussion

In this exploratory study, participants who underwent BWSTT with active or sham TSCS showed heterogeneous, individual-level cardiovascular and autonomic trajectories. Some indices moved in a potentially favourable direction, particularly daytime hypotensive burden, selected HRV measures, stroke volume, and baroreflex engagement (in one TSCS participant), while other outcomes either changed similarly in sham or remained unstable. These findings are of value as “normalizing” cardiovascular control likely requires more than a simple model to increase spinal excitability during ABT. Rather, the cardiovascular response to paired neuromodulation and BWSTT appears variable between individuals and likely influenced by an individual’s autonomic phenotype prior to receiving the intervention.

### Blood pressure regulation and variability

The most clinically relevant pattern was the reduction in the percentage of daytime hypotensive events in the TSCS cohort. This finding is important as persistent low BP and orthostatic intolerance are common after high-level SCI and can directly limit rehabilitation participation.(33) A reduction in hypotensive burden, could therefore be meaningful for daily living.(2) The reduced hypotensive burden from this study aligns with a recently published case-study showing acute improvements in orthostatic tolerance with spinal stimulation, where after six 30-minute stimulation sessions, the participant’s average upright SBP increased from 69 to 88 mmHg and tilt tolerance improved from 3 minutes to the full 30-minute protocol.(34) However, earlier evidence has shown that repeated stand locomotor training can improve orthostatic BP control as 80 sessions had increased seated resting SBP and attenuated the fall in SBP during standing in individuals with motor-complete SCI.(35) In our MACHINE study, one Sham participant showed a decrease in the percentage of daytime hypotensive events. Therefore, the cardiovascular stimulus provided by locomotor training could be augmented by neuromodulation, but because BWSTT was delivered in both groups and the sample was small, we cannot confirm a TSCS-specific effect. Furthermore, the finding should be interpreted alongside the absence of a consistent improvement in overall BP stability. Measures of BPV, including SD-SBP, CoV-SBP, and VIM-SBP, increased more clearly in the Sham cohort overall, although one TSCS participant showed a comparable increase, highlighting inter-individual heterogeneity. Similarly, a small increase in hypertensive (i.e., AD) episodes was observed in both cohorts, although this was less pronounced with TSCS. The presence of these episodes in one Sham participant, together with their absence in Participants 1 (TSCS) and 4 (Sham), suggests that AD may have been influenced by contextual or individual factors rather than by stimulation-specific effects alone. Therefore, the intervention may have influenced the lower daily BP in these participants over the episodic hypertensive instability characteristic of AD. This could be a consideration for future studies as hypotension, orthostatic intolerance, AD frequency, circadian rhythm, and BPV may not respond in parallel and should not be treated as interchangeable markers of “autonomic improvement”.

### Cardiac structure and function

Structural echocardiographic changes were modest; LVIDd remained relatively stable, and the small changes in LVMi were not interpreted as evidence of cardiac remodeling, given the known test-retest variability of echocardiographic LV mass measurements in longitudinal studies.(36) Stability in TSCS participants likely reflects short-term maintenance rather than structural improvement, as no decline occurred in either cohort and meaningful atrophy would be unlikely over three months in chronic SCI. Functional measures diverged more than structural indices. SV responses varied across individuals in both cohorts, while TSCS showed small reductions in E/E’ with minimal E/A change, compatible with reduced filling pressures; Sham demonstrated larger E/E’ reductions alongside increased E/A and LV dimensions, potentially suggesting early diastolic dysfunction rather than normalization. In contrast to our previous CHOICES study, where BWSTT alone did not improve echocardiography outcomes,(26) combining TSCS with BWSTT may provide potential functional benefit in select individuals. These observations align with broader SCI literature showing modest cardiovascular improvements from ABT-based interventions.(35,37)

### Electrocardiography interval variability

TSCS participants showed R-R interval prolongation, modest P-wave and PR lengthening, slight QRS narrowing, and reduced QTc without pathological T_peak_-T_end_ changes (Supplementary Table 1). QTVI also remained negative in both TSCS participants, whereas the Sham cohort showed greater heterogeneity, including one participant with a positive QTVI at Post-Intervention. Because T_peak_-T_end_ and QTVI are markers of ventricular repolarization heterogeneity and temporal repolarization lability, respectively, these findings may suggest relative electrophysiological stability in the TSCS cohort. However, given the small sample size, Pre-Intervention between-participant variability, and the influence of the one Sham participant, this should not be interpreted as evidence of autonomic control, but rather suggest ECG variability metrics may be useful for characterizing individual cardiovascular autonomic profiles in future SCI neuromodulation studies.(38)

### Orthostatic hemodynamics, frequency-domain BPV, HRV and baroreflex

During HUTT, TSCS participants demonstrated smaller changes in TPR, though variability overlapped with Sham. Sham participants appeared to rely on greater peripheral resistance with heterogeneous SV responses, indicating inconsistent SV regulation between cohorts. Similar patterns have been described in case studies where spinal stimulation improved orthostatic tolerance and reduced medication reliance.(34,39) Frequency-domain analyses of BP revealed reduced exaggerated LF oscillations and more organized LF activity in both cohorts after intervention, consistent with autonomic conditioning from repeated BWSTT and orthostatic exposure, as previously suggested from our CHOICES trial.(40) Supine SDNN and RMSSD increased in both cohorts, whereas concurrent increases in LF and HF power were observed only in TSCS (Supplementary Table 1), suggesting broader modulation of cardiac autonomic variability rather than isolated vagal predominance. During HUTT, TSCS participants showed reductions in HF and LF power, consistent with reduced beat-to-beat variability during orthostatic loading (i.e., HUTT), although responses overlapped with Sham. While Sham responses were mixed, baroreflex effectiveness increased in one TSCS participant, primarily through down-sequences, but this could mean that some individuals with chronic motor-complete SCI may retain modifiable cardiovagal-baroreflex coupling, even years after injury. This participant-level response is consistent with prior work showing impaired cardiovagal baroreflex control after SCI and some responsiveness of baroreflex indices to exercise-based interventions,(40) though our findings remain exploratory.

### Translational implications

Electrode placement (T11-L1) was consistent with prior epidural stimulation studies targeting lumbosacral circuits.(41,42) While alignment with established TSCS protocols supports the physiological plausibility of our findings, it does not confirm activation of specific autonomic circuits. Clinical protocols often individualize electrode configurations and stimulation intensity, which may contribute to responder variability. Future trials should incorporate standardized titration protocols and detailed reporting of stimulation exposure (sites, amplitudes, duration, session counts) for reproducibility.(43) Exercise-based interventions in SCI improve some cardiovascular risk markers but generally produce modest changes in BP regulation and autonomic function.(26,40) TSCS may influence spinal autonomic circuitry, however, whether this meaningfully augments rehabilitation-related cardiovascular adaptation remains to be determined in adequately powered trials. In prolonged community-delivered TSCS combined with ABT, clinically meaningful functional changes were not evident by ∼40 sessions but emerged after 60 sessions and continued improving through 120 sessions, suggesting a delayed and cumulative response profile.(44) Autonomic-cardiovascular adaptation may follow a similar trajectory, potentially explaining the modest and heterogeneous changes observed. Future research may explore TSCS combined with other exercise modalities(45) to determine whether cardiovascular adaptation requires a stronger volitional exercise stimulus above the lesion, rather than lower-limb afferent input and orthostatic loading alone. Earlier integration of TSCS into subacute rehabilitation may also be worth exploring as a strategy to mitigate the onset of cardiovascular dysfunction.(46,47)

## Limitations

Although the small number of participants completing the cardiovascular assessments limit our interpretation and prevents reliable between-group conclusions, the study offers a detailed physiological picture of cardiovascular responses to paired neuromodulation and ABT in chronic motor-complete SCI. Pre-Intervention cohort differences in key cardiovascular measures (e.g., median SBP, nocturnal dipping, ECG interval variability), likely related to injury characteristics, were not adjustable, preventing evaluation of TSCS efficacy or dose-response. Training exposure was also individualized, with variation in body-weight support, walking speed, perceived exertion, and walking distance, meaning that the cardiovascular stimulus delivered by BWSTT was not identical across participants. At the same time, the strength of the study lies in showing which cardiovascular measures appear responsive and which remain variable. The sham condition also included BWSTT, and daily life factors may have contributed to ABPM variability. Nevertheless, the combination of ABPM, orthostatic hemodynamics, BPV, echocardiography, ECG-derived indices, HRV, baroreflex analysis, and symptoms provides a useful framework for future targeted trials.

## Conclusion

In this exploratory complete-case analysis, 12 weeks of BWSTT with active or sham TSCS did not produce consistent cardiovascular autonomic improvement across participants. Instead, the results point to more selective, individual-level changes such as fewer daytime hypotensive events with TSCS, reduced symptom burden in both groups, attenuated LF BPV during orthostatic challenge, and evidence of stronger baroreflex engagement in one TSCS participant. Overall, these findings support further investigation of combined BWSTT and TSCS, while suggesting that cardiovascular responses may depend on participants’ autonomic profiles prior to the intervention.

## Data Availability

All data produced in the present study are available upon reasonable request to the authors

## Acknowledgements

The authors thank Dr Parag Gad and SpineX Inc. for the provision of the transcutaneous spinal cord stimulator used in the trial. The authors acknowledge the administrative assistance from Ms. Andrea Maharaj, Ms. Jennifer Phan, Ms. Kawami Cao, Mr. Ali Hosseinzadeh, Dr. Abdullah Alrashidi and Dr. Adam Mesa at various stages of the trial from its conception. Finally, we thank all participants for their commitment and support of this clinical trial.

## Data sharing statement

All data generated and analyzed during this study are included in this published article, further inquiries can be directed to the corresponding author (AVK).

## Ethics statement

The trial and recruitment materials were approved by institutional review board.

## Authors Contributions

SJTB was responsible for analysis and interpretation of the data and original drafting of the manuscript. CS and AMMW were responsible for running the study, collecting, and managing data. SJTB, CS, AMMW, SS, RNM were responsible for data collection. DDH provided technical support with data analysis. AVK conceived the study and is the principal investigator who managed data collection. The study was conceived with expert support and input from SJTB, AMMW, TEN, RS, SLE, MJB, TL, and AVK. All authors provided critical revision of the manuscript and final approval of the version to be published.

---

**Supplementary Table 1:**
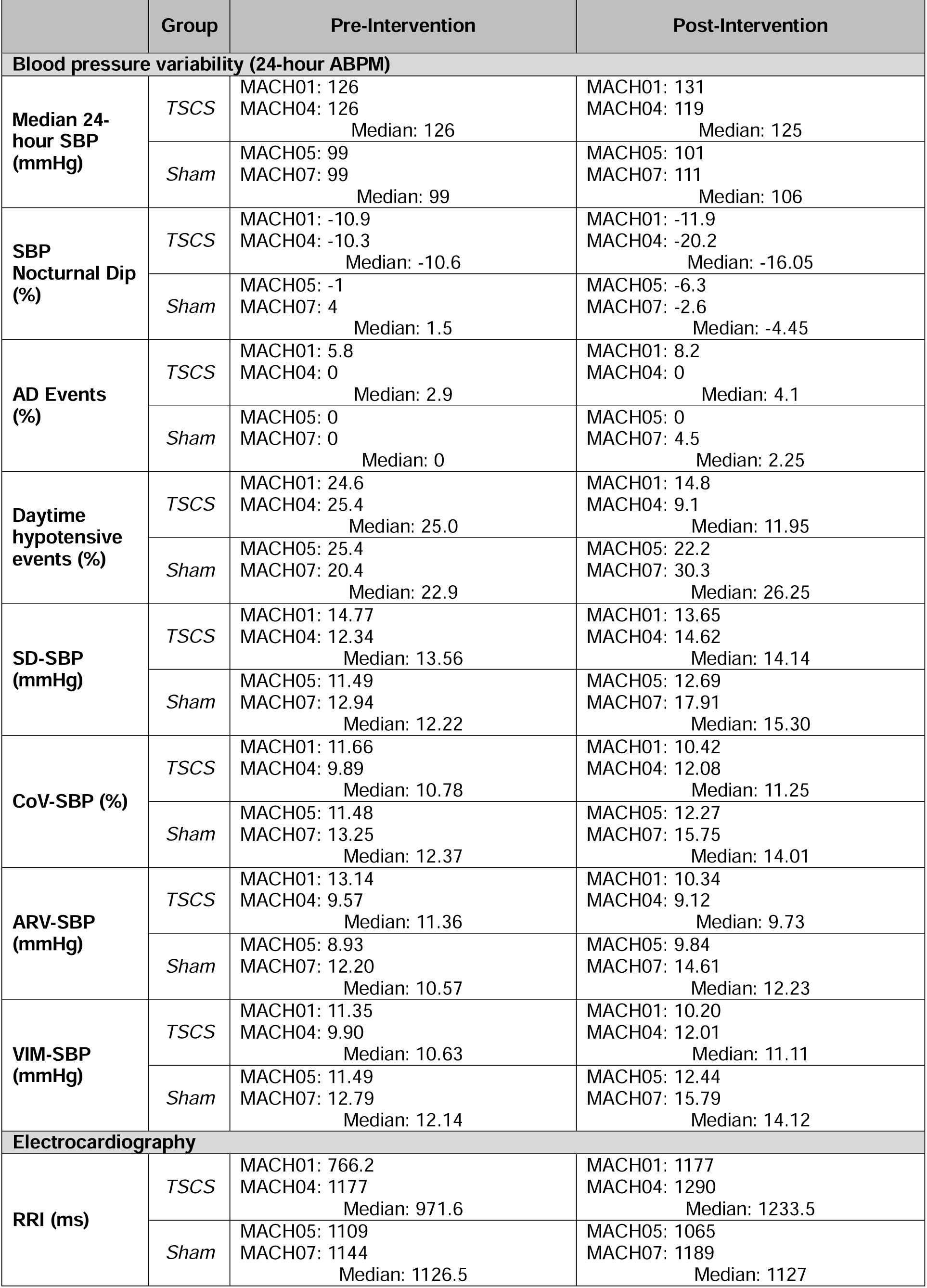
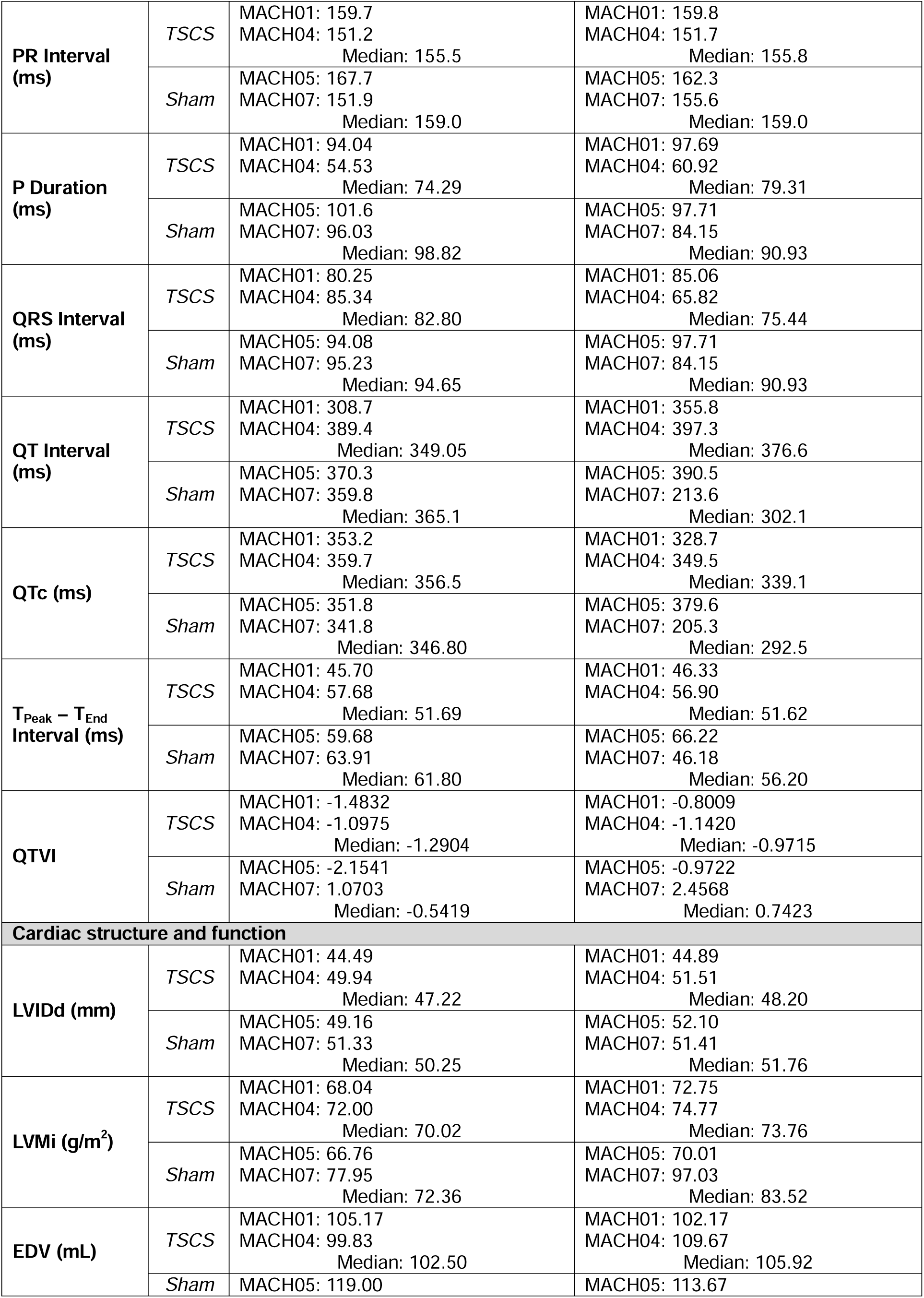
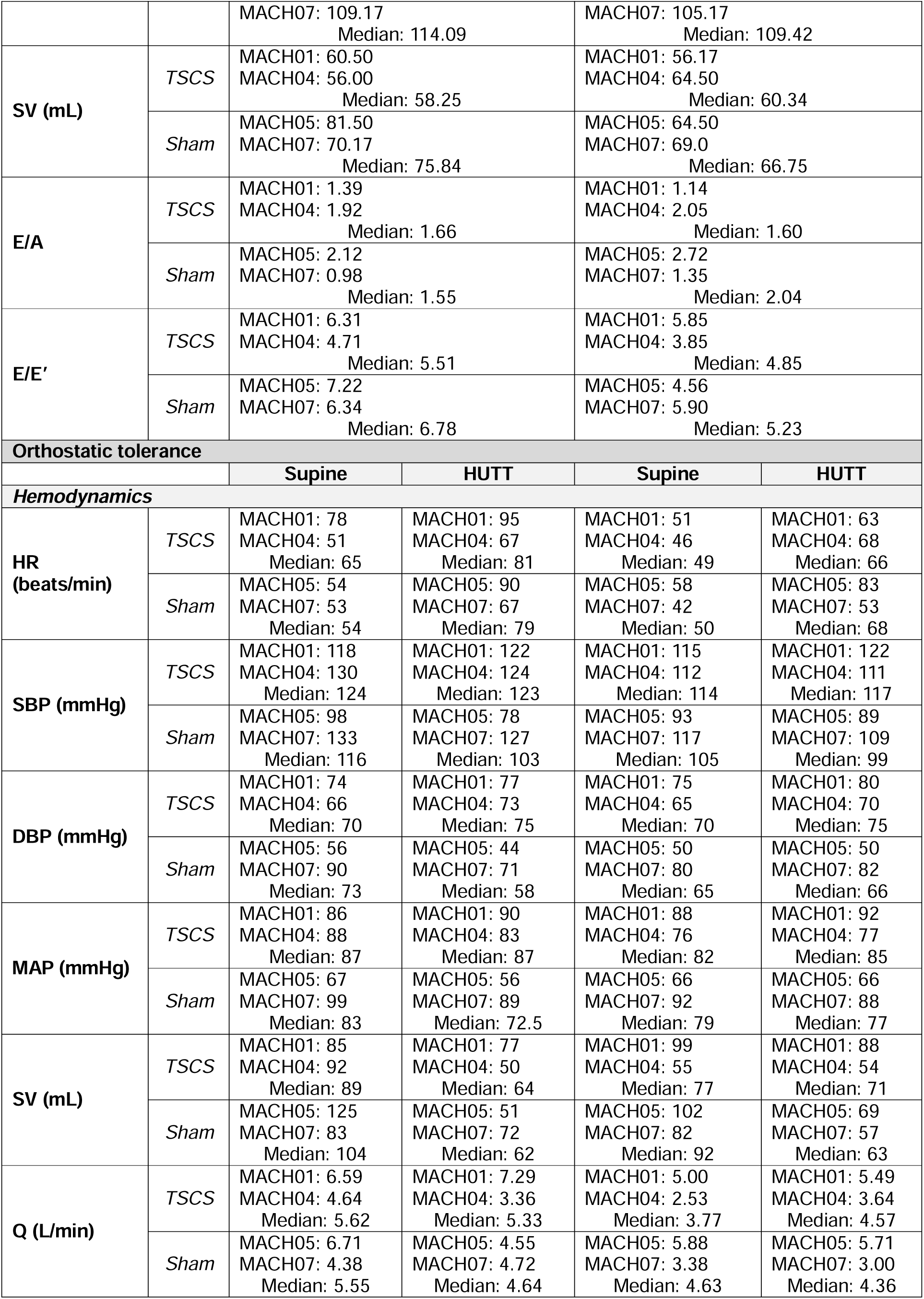
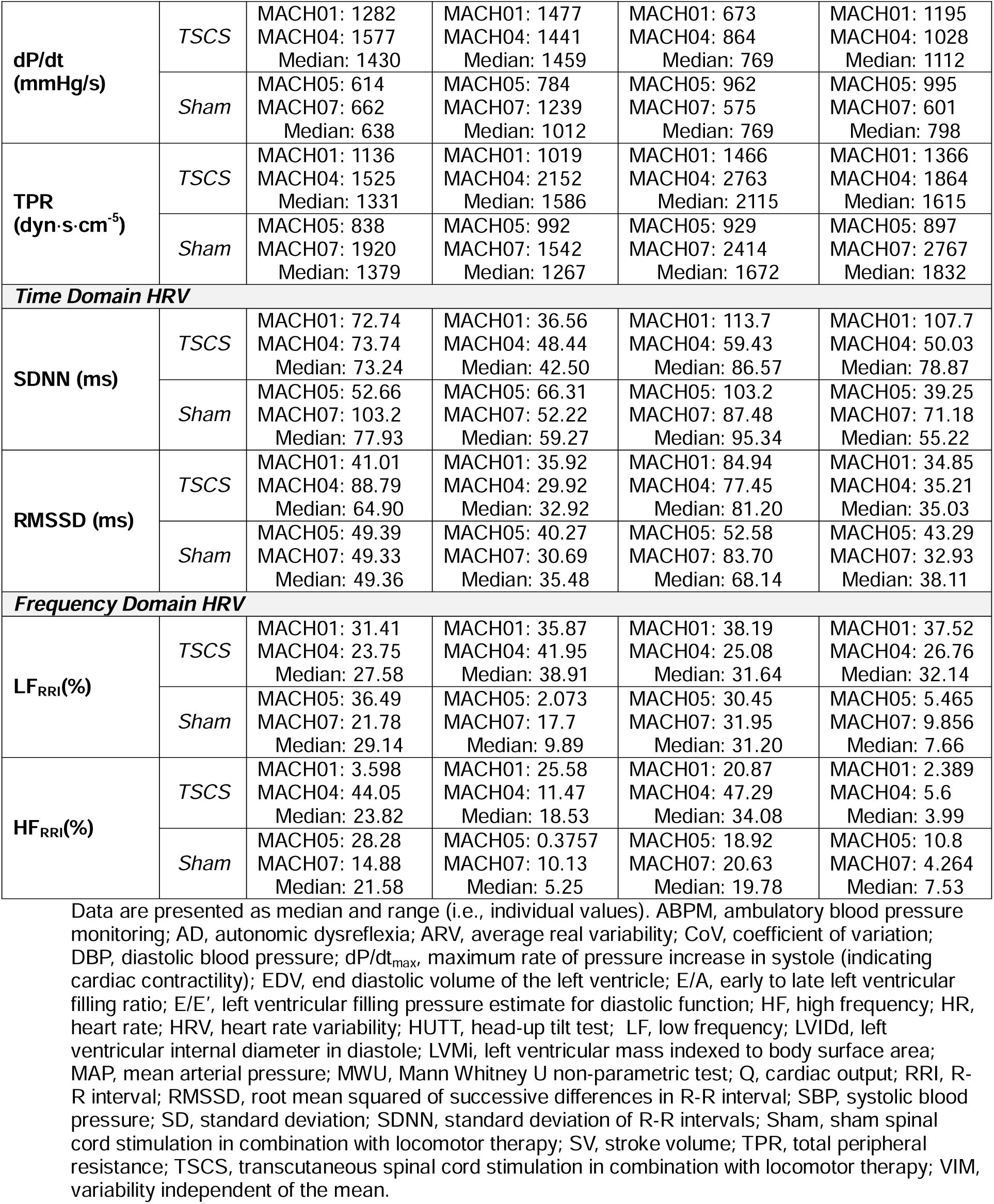

|  | Group | Pre-Intervention | Post-Intervention |
| --- | --- | --- | --- |
| **Blood pressure variability (24-hour ABPM)** |  |  |  |
| **Median 24-hour SBP (mmHg)** | *TSCS* | MACH01: 126<br>MACH04: 126<br>Median: 126 | MACH01: 131<br>MACH04: 119<br>Median: 125 |
|  | *Sham* | MACH05: 99<br>MACH07: 99<br>Median: 99 | MACH05: 101<br>MACH07: 111<br>Median: 106 |
| **SBP Nocturnal Dip (%)** | *TSCS* | MACH01: -10.9<br>MACH04: -10.3<br>Median: -10.6 | MACH01: -11.9<br>MACH04: -20.2<br>Median: -16.05 |
|  | *Sham* | MACH05: -1<br>MACH07: 4<br>Median: 1.5 | MACH05: -6.3<br>MACH07: -2.6<br>Median: -4.45 |
| **AD Events (%)** | *TSCS* | MACH01: 5.8<br>MACH04: 0<br>Median: 2.9 | MACH01: 8.2<br>MACH04: 0<br>Median: 4.1 |
|  | *Sham* | MACH05: 0<br>MACH07: 0<br>Median: 0 | MACH05: 0<br>MACH07: 4.5<br>Median: 2.25 |
| **Daytime hypotensive events (%)** | *TSCS* | MACH01: 24.6<br>MACH04: 25.4<br>Median: 25.0 | MACH01: 14.8<br>MACH04: 9.1<br>Median: 11.95 |
|  | *Sham* | MACH05: 25.4<br>MACH07: 20.4<br>Median: 22.9 | MACH05: 22.2<br>MACH07: 30.3<br>Median: 26.25 |
| **SD-SBP (mmHg)** | *TSCS* | MACH01: 14.77<br>MACH04: 12.34<br>Median: 13.56 | MACH01: 13.65<br>MACH04: 14.62<br>Median: 14.14 |
|  | *Sham* | MACH05: 11.49<br>MACH07: 12.94<br>Median: 12.22 | MACH05: 12.69<br>MACH07: 17.91<br>Median: 15.30 |
| **CoV-SBP (%)** | *TSCS* | MACH01: 11.66<br>MACH04: 9.89<br>Median: 10.78 | MACH01: 10.42<br>MACH04: 12.08<br>Median: 11.25 |
|  | *Sham* | MACH05: 11.48<br>MACH07: 13.25<br>Median: 12.37 | MACH05: 12.27<br>MACH07: 15.75<br>Median: 14.01 |
| **ARV-SBP (mmHg)** | *TSCS* | MACH01: 13.14<br>MACH04: 9.57<br>Median: 11.36 | MACH01: 10.34<br>MACH04: 9.12<br>Median: 9.73 |
|  | *Sham* | MACH05: 8.93<br>MACH07: 12.20<br>Median: 10.57 | MACH05: 9.84<br>MACH07: 14.61<br>Median: 12.23 |
| **VIM-SBP (mmHg)** | *TSCS* | MACH01: 11.35<br>MACH04: 9.90<br>Median: 10.63 | MACH01: 10.20<br>MACH04: 12.01<br>Median: 11.11 |
|  | *Sham* | MACH05: 11.49<br>MACH07: 12.79<br>Median: 12.14 | MACH05: 12.44<br>MACH07: 15.79<br>Median: 14.12 |
| **Electrocardiography** |  |  |  |
| **RRI (ms)** | *TSCS* | MACH01: 766.2<br>MACH04: 1177<br>Median: 971.6 | MACH01: 1177<br>MACH04: 1290<br>Median: 1233.5 |
|  | *Sham* | MACH05: 1109<br>MACH07: 1144<br>Median: 1126.5 | MACH05: 1065<br>MACH07: 1189<br>Median: 1127 |
| **PR Interval (ms)** | *TSCS* | MACH01: 159.7<br>MACH04: 151.2<br>Median: 155.5 | MACH01: 159.8<br>MACH04: 151.7<br>Median: 155.8 |
|  | *Sham* | MACH05: 167.7<br>MACH07: 151.9<br>Median: 159.0 | MACH05: 162.3<br>MACH07: 155.6<br>Median: 159.0 |
| **P Duration (ms)** | *TSCS* | MACH01: 94.04<br>MACH04: 54.53<br>Median: 74.29 | MACH01: 97.69<br>MACH04: 60.92<br>Median: 79.31 |
|  | *Sham* | MACH05: 101.6<br>MACH07: 96.03<br>Median: 98.82 | MACH05: 97.71<br>MACH07: 84.15<br>Median: 90.93 |
| **QRS Interval (ms)** | *TSCS* | MACH01: 80.25<br>MACH04: 85.34<br>Median: 82.80 | MACH01: 85.06<br>MACH04: 65.82<br>Median: 75.44 |
|  | *Sham* | MACH05: 94.08<br>MACH07: 95.23<br>Median: 94.65 | MACH05: 97.71<br>MACH07: 84.15<br>Median: 90.93 |
| **QT Interval (ms)** | *TSCS* | MACH01: 308.7<br>MACH04: 389.4<br>Median: 349.05 | MACH01: 355.8<br>MACH04: 397.3<br>Median: 376.6 |
|  | *Sham* | MACH05: 370.3<br>MACH07: 359.8<br>Median: 365.1 | MACH05: 390.5<br>MACH07: 213.6<br>Median: 302.1 |
| **QTc (ms)** | *TSCS* | MACH01: 353.2<br>MACH04: 359.7<br>Median: 356.5 | MACH01: 328.7<br>MACH04: 349.5<br>Median: 339.1 |
|  | *Sham* | MACH05: 351.8<br>MACH07: 341.8<br>Median: 346.80 | MACH05: 379.6<br>MACH07: 205.3<br>Median: 292.5 |
| **T<sub>Peak</sub> – T<sub>End</sub> Interval (ms)** | *TSCS* | MACH01: 45.70<br>MACH04: 57.68<br>Median: 51.69 | MACH01: 46.33<br>MACH04: 56.90<br>Median: 51.62 |
|  | *Sham* | MACH05: 59.68<br>MACH07: 63.91<br>Median: 61.80 | MACH05: 66.22<br>MACH07: 46.18<br>Median: 56.20 |
| **QTVI** | *TSCS* | MACH01: -1.4832<br>MACH04: -1.0975<br>Median: -1.2904 | MACH01: -0.8009<br>MACH04: -1.1420<br>Median: -0.9715 |
|  | *Sham* | MACH05: -2.1541<br>MACH07: 1.0703<br>Median: -0.5419 | MACH05: -0.9722<br>MACH07: 2.4568<br>Median: 0.7423 |
| **Cardiac structure and function** |  |  |  |
| **LVIDd (mm)** | *TSCS* | MACH01: 44.49<br>MACH04: 49.94<br>Median: 47.22 | MACH01: 44.89<br>MACH04: 51.51<br>Median: 48.20 |
|  | *Sham* | MACH05: 49.16<br>MACH07: 51.33<br>Median: 50.25 | MACH05: 52.10<br>MACH07: 51.41<br>Median: 51.76 |
| **LVMi (g/m<sup>2</sup>)** | *TSCS* | MACH01: 68.04<br>MACH04: 72.00<br>Median: 70.02 | MACH01: 72.75<br>MACH04: 74.77<br>Median: 73.76 |
|  | *Sham* | MACH05: 66.76<br>MACH07: 77.95<br>Median: 72.36 | MACH05: 70.01<br>MACH07: 97.03<br>Median: 83.52 |
| **EDV (mL)** | *TSCS* | MACH01: 105.17<br>MACH04: 99.83<br>Median: 102.50 | MACH01: 102.17<br>MACH04: 109.67<br>Median: 105.92 |
|  | *Sham* | MACH05: 119.00 | MACH05: 113.67 |

|  |  |  |  |  |  |
| --- | --- | --- | --- | --- | --- |
|  |  | MACH07: 109.17<br>Median: 114.09 |  | MACH07: 105.17<br>Median: 109.42 |  |
| SV (mL) | TSCS | MACH01: 60.50<br>MACH04: 56.00<br>Median: 58.25 |  | MACH01: 56.17<br>MACH04: 64.50<br>Median: 60.34 |  |
|  | Sham | MACH05: 81.50<br>MACH07: 70.17<br>Median: 75.84 |  | MACH05: 64.50<br>MACH07: 69.0<br>Median: 66.75 |  |
| E/A | TSCS | MACH01: 1.39<br>MACH04: 1.92<br>Median: 1.66 |  | MACH01: 1.14<br>MACH04: 2.05<br>Median: 1.60 |  |
|  | Sham | MACH05: 2.12<br>MACH07: 0.98<br>Median: 1.55 |  | MACH05: 2.72<br>MACH07: 1.35<br>Median: 2.04 |  |
| E/E' | TSCS | MACH01: 6.31<br>MACH04: 4.71<br>Median: 5.51 |  | MACH01: 5.85<br>MACH04: 3.85<br>Median: 4.85 |  |
|  | Sham | MACH05: 7.22<br>MACH07: 6.34<br>Median: 6.78 |  | MACH05: 4.56<br>MACH07: 5.90<br>Median: 5.23 |  |
| Orthostatic tolerance |  |  |  |  |  |
|  |  | Supine | HUTT | Supine | HUTT |
| Hemodynamics |  |  |  |  |  |
| HR (beats/min) | TSCS | MACH01: 78<br>MACH04: 51<br>Median: 65 | MACH01: 95<br>MACH04: 67<br>Median: 81 | MACH01: 51<br>MACH04: 46<br>Median: 49 | MACH01: 63<br>MACH04: 68<br>Median: 66 |
|  | Sham | MACH05: 54<br>MACH07: 53<br>Median: 54 | MACH05: 90<br>MACH07: 67<br>Median: 79 | MACH05: 58<br>MACH07: 42<br>Median: 50 | MACH05: 83<br>MACH07: 53<br>Median: 68 |
| SBP (mmHg) | TSCS | MACH01: 118<br>MACH04: 130<br>Median: 124 | MACH01: 122<br>MACH04: 124<br>Median: 123 | MACH01: 115<br>MACH04: 112<br>Median: 114 | MACH01: 122<br>MACH04: 111<br>Median: 117 |
|  | Sham | MACH05: 98<br>MACH07: 133<br>Median: 116 | MACH05: 78<br>MACH07: 127<br>Median: 103 | MACH05: 93<br>MACH07: 117<br>Median: 105 | MACH05: 89<br>MACH07: 109<br>Median: 99 |
| DBP (mmHg) | TSCS | MACH01: 74<br>MACH04: 66<br>Median: 70 | MACH01: 77<br>MACH04: 73<br>Median: 75 | MACH01: 75<br>MACH04: 65<br>Median: 70 | MACH01: 80<br>MACH04: 70<br>Median: 75 |
|  | Sham | MACH05: 56<br>MACH07: 90<br>Median: 73 | MACH05: 44<br>MACH07: 71<br>Median: 58 | MACH05: 50<br>MACH07: 80<br>Median: 65 | MACH05: 50<br>MACH07: 82<br>Median: 66 |
| MAP (mmHg) | TSCS | MACH01: 86<br>MACH04: 88<br>Median: 87 | MACH01: 90<br>MACH04: 83<br>Median: 87 | MACH01: 88<br>MACH04: 76<br>Median: 82 | MACH01: 92<br>MACH04: 77<br>Median: 85 |
|  | Sham | MACH05: 67<br>MACH07: 99<br>Median: 83 | MACH05: 56<br>MACH07: 89<br>Median: 72.5 | MACH05: 66<br>MACH07: 92<br>Median: 79 | MACH05: 66<br>MACH07: 88<br>Median: 77 |
| SV (mL) | TSCS | MACH01: 85<br>MACH04: 92<br>Median: 89 | MACH01: 77<br>MACH04: 50<br>Median: 64 | MACH01: 99<br>MACH04: 55<br>Median: 77 | MACH01: 88<br>MACH04: 54<br>Median: 71 |
|  | Sham | MACH05: 125<br>MACH07: 83<br>Median: 104 | MACH05: 51<br>MACH07: 72<br>Median: 62 | MACH05: 102<br>MACH07: 82<br>Median: 92 | MACH05: 69<br>MACH07: 57<br>Median: 63 |
| Q (L/min) | TSCS | MACH01: 6.59<br>MACH04: 4.64<br>Median: 5.62 | MACH01: 7.29<br>MACH04: 3.36<br>Median: 5.33 | MACH01: 5.00<br>MACH04: 2.53<br>Median: 3.77 | MACH01: 5.49<br>MACH04: 3.64<br>Median: 4.57 |
|  | Sham | MACH05: 6.71<br>MACH07: 4.38<br>Median: 5.55 | MACH05: 4.55<br>MACH07: 4.72<br>Median: 4.64 | MACH05: 5.88<br>MACH07: 3.38<br>Median: 4.63 | MACH05: 5.71<br>MACH07: 3.00<br>Median: 4.36 |
| **dP/dt**<br>**(mmHg/s)** | *TSCS* | MACH01: 1282<br>MACH04: 1577<br>Median: 1430 | MACH01: 1477<br>MACH04: 1441<br>Median: 1459 | MACH01: 673<br>MACH04: 864<br>Median: 769 | MACH01: 1195<br>MACH04: 1028<br>Median: 1112 |
|  | *Sham* | MACH05: 614<br>MACH07: 662<br>Median: 638 | MACH05: 784<br>MACH07: 1239<br>Median: 1012 | MACH05: 962<br>MACH07: 575<br>Median: 769 | MACH05: 995<br>MACH07: 601<br>Median: 798 |
| **TPR**<br>**(dyn·s·cm<sup>-5</sup>)** | *TSCS* | MACH01: 1136<br>MACH04: 1525<br>Median: 1331 | MACH01: 1019<br>MACH04: 2152<br>Median: 1586 | MACH01: 1466<br>MACH04: 2763<br>Median: 2115 | MACH01: 1366<br>MACH04: 1864<br>Median: 1615 |
|  | *Sham* | MACH05: 838<br>MACH07: 1920<br>Median: 1379 | MACH05: 992<br>MACH07: 1542<br>Median: 1267 | MACH05: 929<br>MACH07: 2414<br>Median: 1672 | MACH05: 897<br>MACH07: 2767<br>Median: 1832 |
| **Time Domain HRV** |  |  |  |  |  |
| **SDNN (ms)** | *TSCS* | MACH01: 72.74<br>MACH04: 73.74<br>Median: 73.24 | MACH01: 36.56<br>MACH04: 48.44<br>Median: 42.50 | MACH01: 113.7<br>MACH04: 59.43<br>Median: 86.57 | MACH01: 107.7<br>MACH04: 50.03<br>Median: 78.87 |
|  | *Sham* | MACH05: 52.66<br>MACH07: 103.2<br>Median: 77.93 | MACH05: 66.31<br>MACH07: 52.22<br>Median: 59.27 | MACH05: 103.2<br>MACH07: 87.48<br>Median: 95.34 | MACH05: 39.25<br>MACH07: 71.18<br>Median: 55.22 |
| **RMSSD (ms)** | *TSCS* | MACH01: 41.01<br>MACH04: 88.79<br>Median: 64.90 | MACH01: 35.92<br>MACH04: 29.92<br>Median: 32.92 | MACH01: 84.94<br>MACH04: 77.45<br>Median: 81.20 | MACH01: 34.85<br>MACH04: 35.21<br>Median: 35.03 |
|  | *Sham* | MACH05: 49.39<br>MACH07: 49.33<br>Median: 49.36 | MACH05: 40.27<br>MACH07: 30.69<br>Median: 35.48 | MACH05: 52.58<br>MACH07: 83.70<br>Median: 68.14 | MACH05: 43.29<br>MACH07: 32.93<br>Median: 38.11 |
| **Frequency Domain HRV** |  |  |  |  |  |
| **LF<sub>RRI</sub>(%)** | *TSCS* | MACH01: 31.41<br>MACH04: 23.75<br>Median: 27.58 | MACH01: 35.87<br>MACH04: 41.95<br>Median: 38.91 | MACH01: 38.19<br>MACH04: 25.08<br>Median: 31.64 | MACH01: 37.52<br>MACH04: 26.76<br>Median: 32.14 |
|  | *Sham* | MACH05: 36.49<br>MACH07: 21.78<br>Median: 29.14 | MACH05: 2.073<br>MACH07: 17.7<br>Median: 9.89 | MACH05: 30.45<br>MACH07: 31.95<br>Median: 31.20 | MACH05: 5.465<br>MACH07: 9.856<br>Median: 7.66 |
| **HF<sub>RRI</sub>(%)** | *TSCS* | MACH01: 3.598<br>MACH04: 44.05<br>Median: 23.82 | MACH01: 25.58<br>MACH04: 11.47<br>Median: 18.53 | MACH01: 20.87<br>MACH04: 47.29<br>Median: 34.08 | MACH01: 2.389<br>MACH04: 5.6<br>Median: 3.99 |
|  | *Sham* | MACH05: 28.28<br>MACH07: 14.88<br>Median: 21.58 | MACH05: 0.3757<br>MACH07: 10.13<br>Median: 5.25 | MACH05: 18.92<br>MACH07: 20.63<br>Median: 19.78 | MACH05: 10.8<br>MACH07: 4.264<br>Median: 7.53 |

Data are presented as median and range (i.e., individual values). ABPM, ambulatory blood pressure monitoring; AD, autonomic dysreflexia; ARV, average real variability; CoV, coefficient of variation; DBP, diastolic blood pressure; dP/dt<sub>max</sub>, maximum rate of pressure increase in systole (indicating cardiac contractility); EDV, end diastolic volume of the left ventricle; E/A, early to late left ventricular filling ratio; E/E', left ventricular filling pressure estimate for diastolic function; HF, high frequency; HR, heart rate; HRV, heart rate variability; HUTT, head-up tilt test; LF, low frequency; LVIDd, left ventricular internal diameter in diastole; LVMI, left ventricular mass indexed to body surface area; MAP, mean arterial pressure; MWU, Mann Whitney U non-parametric test; Q, cardiac output; RRI, R-R interval; RMSSD, root mean squared of successive differences in R-R interval; SBP, systolic blood pressure; SD, standard deviation; SDNN, standard deviation of R-R intervals; Sham, sham spinal cord stimulation in combination with locomotor therapy; SV, stroke volume; TPR, total peripheral resistance; TSCS, transcutaneous spinal cord stimulation in combination with locomotor therapy; VIM, variability independent of the mean.

